# A critical view of the use of predictive energy equations for the identification of hypermetabolism in motor neuron disease

**DOI:** 10.1101/2022.12.19.22283673

**Authors:** Sarah Roscoe, Ellie Skinner, Elaine Kabucho Kibirige, Charmaine Childs, C. Elizabeth Weekes, Stephen Wootton, Scott Allen, Christopher McDermott, Theocharis Stavroulakis

**Affiliations:** Sheffield Institute for Translational Neuroscience, The University of Sheffield, Sheffield, UK; College of Health, Wellbeing and Life Sciences, Sheffield Hallam University, Sheffield, UK; Department of Nutrition & Dietetics, Guy’s & St Thomas’ NHS Foundation Trust, London, UK; Faculty of Medicine, University of Southampton, Southampton, UK; Southampton NIHR Biomedical Research Centre, University Hospital Southampton, Southampton, UK

**Author notes:** Equal contribution (Scott Allen, Christopher McDermott, Theocharis Stavroulakis). **Author emails:** Sarah Roscoe, Ellie Skinner, Elaine Kabucho Kibirige, Charmaine Childs, C. Elizabeth Weekes, Stephen Wootton Scott Allen, Christopher McDermott, Theocharis Stavroulakis.

**Keywords:** Motor neuron(e) disease, Hypermetabolism, Malnutrition, Resting Energy Expenditure, Indirect Calorimetry, Predictive Energy Equations

## Abstract

**Background and Aims:** People living with motor neuron disease (MND) frequently struggle to consume an optimal caloric intake. Often compounded by hypermetabolism, this can lead to dysregulated energy homeostasis, prompting the onset of malnutrition and associated weight loss. This is associated with a poorer prognosis and reduced survival. It is therefore important to establish appropriate nutritional goals to ensure adequate energy intake. This is best done by measuring resting energy expenditure (mREE) using indirect calorimetry. However, indirect calorimetry is not widely available in clinical practice, thus dietitians caring for people living with MND frequently use energy equations to predict resting energy expenditure (pREE) and estimate caloric requirements. Energy prediction equations have previously been shown to underestimate resting energy expenditure in over two-thirds of people living with MND.

Hypermetabolism has previously been identified using the metabolic index. The metabolic index is a ratio of mREE to pREE, whereby an increase of mREE by ≥ 110% indicates hypermetabolism. We propose that the use of energy prediction equations to inform a metabolic index to indicate hypermetabolism in people living with MND is inappropriate and results in a biased identification of hypermetabolism in lighter individuals.

**Methods:** mREE was derived using VO_2_ and VCO_2_ measurements from a GEMNutrition indirect calorimeter. pREE was estimated by Harris-Benedict (HB) (1919), Henry (2005) and kcal/kg/day predictive energy equations. The REE variation, described as the percentage difference between mREE and pREE, determined the accuracy of pREE ([pREE-mREE]/mREE) x 100), with accuracy defined as ≤ ± 10%. A metabolic index threshold of ≥ 110% was used to classify hypermetabolism. All resting energy expenditure data are presented as kcal/24hr.

**Results:** Sixteen people living with MND were included in the analysis. The mean mREE was 1642 kcal/24hr ranging between 1110 and 2015 kcal/24hr. When REE variation was analysed for the entire cohort, the HB, Henry and kcal/kg/day equations all overestimated REE, but remained within the accuracy threshold (mean values were 2.81% for HB, 4.51% for Henry and 8.00% for kcal/kg/day). Conversely, inter-individual REE variation within the cohort revealed HB and Henry equations both inaccurately reflected mREE for 68.7% of participants, with kcal/kg/day inaccurately reflecting 41.7% of participants. Whilst the overall cohort was not classified as hypermetabolic (mean values were 101.04% for HB, 98.62% for Henry and 95.64% for kcal/kg/day), the metabolic index ranges within the cohort were 70.75% - 141.58% for HB, 72.82% - 127.69% for Henry and 66.09% – 131.58% for kcal/kg/day, indicating both over- and under-estimation of REE by these equations. We have shown that pREE correlates with body weight (kg), whereby the lighter the individual, the greater the underprediction of REE. When applied to the metabolic index, this underprediction biases towards the classification of hypermetabolism in lighter individuals.

**Conclusion:** Whilst predicting resting energy expenditure using the HB, Henry or kcal/kg/day equations accurately reflects derived mREE at group level, these equations are not suitable for informing resting energy expenditure and classification of hypermetabolism when applied to individuals in clinical practice.

## Introduction

Motor neuron disease (MND) encompasses an incurable heterogeneous group of progressive neurodegenerative motor syndromes involving the gradual degeneration and ultimate death of motor neurons. This leads to the weakness and wasting of muscles controlling movement, speech and breathing (1), resulting in death typically from respiratory failure approximately two-to-three years post diagnosis (2,3). The prevalence of MND is 3.37 per 100,000 people worldwide (4) with Amyotrophic Lateral Sclerosis (ALS), the most common form of MND, comprising an estimated 65-85% of cases (5).

Weight loss in people living with MND (plwMND) is primarily driven by the relentless progression of denervation-induced muscle wasting. Symptoms such as dysphagia and a decreased dexterity secondary to muscle weakness (6–8), contribute to a sub-optimal caloric intake, which may lead to malnutrition and further weight loss (9–12). The presence of hypermetabolism, i.e., the state of an increased resting energy expenditure (REE), can result in dysregulated energy homeostasis and thus exacerbate the nutritional challenges for plwMND (13). Individuals with the greatest energy imbalance exhibit a faster rate of functional decline and shorter survival (9,14–18).

It is therefore important to accurately estimate an individual’s total daily energy expenditure (TDEE) to establish appropriate nutritional energy intake goals. REE, i.e., the amount of energy required to maintain normal physiology at rest (19), comprises 60% of TDEE, the remainder of which is exerted through physical activity and the thermic effect from food metabolism (20). REE is best calculated using indirect calorimetry, which directly measures inspired O_2_ and expired CO_2_ to derive measures of REE (mREE). However, indirect calorimetry may be costly, time consuming and not readily available in all clinical contexts (21). When it is not possible to perform indirect calorimetry, REE is predicted (pREE) using predictive energy equations (22). The Henry equation (23) is reported to be the most commonly utilised predictive energy equation by dietitians caring for plwMND in the UK (22).

The assessment for the presence of hypermetabolism involves the calculation of the metabolic index, i.e., the ratio of mREE to pREE, expressed as a percentage. It is accepted that a metabolic index of ≥ 110% typically indicates hypermetabolism (9,24–30). The Harris-Benedict (HB) (1919) (31) predictive equation is frequently used as the denominator in the metabolic index calculation (8,32–36). This is despite the discouragement of the use of the HB equation in MND clinical care in the UK, as it may poorly reflect REE in approximately half of cases (37,38). Nonetheless, application of the metabolic index using the HB equation has previously indicated that 50-68% of plwMND are considered hypermetabolic (24–26,28,29).

We aim to critically reflect on the current unchallenged use of predictive energy equations as comparators against mREE to calculate the metabolic index in plwMND. To achieve this, we focus primarily on two commonly used predictive energy equations in MND; the Henry (23,39) and HB (31). We have also evaluated the suitability of pREE using kcal/kg/day (40) in our cohort.

## Material and methods

### Participant recruitment

Twenty-four plwMND were recruited from the Sheffield MND Care and Research Centre, Royal Hallamshire Hospital, Sheffield Teaching Hospitals NHS Foundation Trust, from October 2021 to August 2022. Favourable opinion for this research was obtained from the London-Fulham Research Ethics Committee 21/PR/0092.

### Inclusion criteria

Participants included with a confirmed diagnosis of MND were invited to participate. Time since diagnosis, MND phenotype, site of onset and medication were not considered for eligibility. Exclusion criteria were limited to an underlying, unmanaged significant co-morbidity that would affect survival or metabolic state, independent of MND (e.g., thyroid disease, cancer), or significant decision-making incapacity preventing informed consent.

### Data collection

This study presents cross-sectional data from baseline visits collected during a longitudinal, observational, prospective study. Study visits were conducted at the Advanced Wellbeing Research Centre, Sheffield Hallam University. The following information was collected from each participant, where possible: demographic; clinical; anthropometric; indirect calorimetry; 24hr urinary collections.

### Anthropometric measurements

Weight and height measurements were recorded in light clothing and shoes in an unaided standing position. Participant-reported weight and height measurements were collected from participants unable to stand unaided for those that could recall a recent measurement. BMI (kg/m^2^) was calculated using: BMI (kg/m^2^) = weight (kg)/height^2^ (m). Arm muscle area (AMA) was calculated using the triceps skinfold (TSF) and mid-upper arm circumference (MUAC) values for the left and right arms: AMA (cm^2^) = [MUAC – (TSF x Π)]^2^ / (4 x Π) as a proxy for lean body mass (LBM).

### Total urinary nitrogen

Total urinary nitrogen (TUN) (g/24hr) was measured from 24-hour urinary collections following Micro-Kjeldahl analysis (41,42). To ensure adherence to the provision of a complete 24-hour urinary collection, participants were requested to record the start and end timings of their collection, as well as timings of all samples collected and details of any spillages or missed collections. Samples were deemed complete if collected over the appropriate 24-hour period and no missed collections or spillages. Incomplete collections were not included in analysis.

### Measured resting energy expenditure

mREE in kcal/24hr was derived following indirect calorimetry using the GEMNutrition Gas Exchange Measurement (GEM) open-circuit metabolic cart with canopy hood. The GEM was calibrated using Laserpure nitrogen and 1% CO_2_/20 % O_2_/N_2_ calibration gases. A realistic, pragmatic approach was adopted to conduct indirect calorimetry and derive mREE in this cohort. Participants were rested in a seated position for one hour prior to measurement. Calorimetry measurement lasted 20 minutes in either a semi-supine or seated position, allowing for participant mobility and respiratory complications. The time of day for the calorimetry measurement was not standardised, but instead influenced by participant and carer availability to reduce burden; participants were therefore not required to be in a fasted state. Participants did not sleep or talk during the measurement. The first five minutes of measurements were discounted from analysis to increase the possibility of reaching a steady state (coefficient of variation (CV) ≤ 5%).

Fractional measures of inspired (Fi) and expired (Fe) O_2_ and CO_2_ measured directly by the GEM were derived into VO₂ and VCO₂ measurements using Haldane’s transformation (43). Measures of VO2, VCO₂and TUN were then applied to the Weir equation to derive the mREE: mREE = ((3.941 x VO_2_) + (1.106 x VCO_2_)) x 1.44 -(2.17 x TUN) (44). The inclusion of total urinary nitrogen in the Weir equation reduces measurement error to provide the most accurate derivation of mREE possible.

### Predicted energy expenditure

pREE was estimated in kcal/24hrs by the HB (1919) (31) and Henry (2005) (23) energy prediction equations (Table 1). Kcal per kg body weight per day (kcal/kg/day) was also calculated based on body weight; i.e., 22 kcal/kg/day was applied to those ≤ 65 years of age, and 24 kcal/kg/day to those > 65 years (40). Participants with BMI values ≤ 18.5 or ≥ 30.0 kg/m^2^ were excluded from analysis (n = 4 (25% of the original cohort)).

**Table 1:** Harris-Benedict (1919) and Henry (2005) predictive energy equations according to sex and age group.

|  | Sex | Age | Predictive energy equation |
| --- | --- | --- | --- |
| **Harris-Benedict** | **Male** | **N/A** | $66.47 + (13.75 \times \text{weight (kg)}) + (5.0 \times \text{height (cm)}) - (6.75 \times \text{age (years)})$ |
| | **Female** | | $655.09 + (9.56 \times \text{weight (kg)}) + (1.84 \times \text{height (cm)}) - (4.67 \times \text{age (years)})$ |
| **Henry** | **Male** | **18-30** | $(14.4 \times \text{weight (kg)} + (313 \times \text{height (m)}) + 113$ |
| | | **30-60** | $(11.4 \times \text{weight (kg)} + (541 \times \text{height (m)}) - 13$ |
| | | **60+** | $(11.4 \times \text{weight (kg)} + (541 \times \text{height (m)}) - 256$ |
| | **Female** | **18-30** | $(10.4 \times \text{weight (kg)} + (615 \times \text{height (m)}) - 282$ |
| | | **30-60** | $(8.18 \times \text{weight (kg)} + (502 \times \text{height (m)}) - 11.6$ |
| | | **60+** | $(8.52 \times \text{weight (kg)} + (421 \times \text{height (m)}) + 10.7$ |

### Statistical analysis

Statistical analysis was conducted using IBM^®^ SPSS^®^ Statistics v27 and GraphPad Prism v9.3.1 (GraphPad Software Inc, La Jolla, CA, USA). Continuous variables were presented as mean ± one standard deviation (SD). Reported kcal/24hr were rounded to the nearest whole number. Mean values were compared using dependent t-tests. Normality was assessed using the Shapiro-Wilk test. Spearman or Pearson bivariate correlation analysis was performed according to the results from the Shapiro-Wilk test. Correlations were plotted with a linear regression line and 95 % confidence intervals from the mean. The threshold for significance was *p* ≤0.05 for all analyses. Bland-Altman limits of agreement analysis (mean bias ± 95% confidence intervals) was used to assess the extent of error of each predictive equation by comparison against mREE (45). Mean bias demonstrates the average difference between measured and predicted REE at group level.

### REE Variation

The REE variation, i.e., the percentage difference between pREE and mREE (%ΔREE), to determine the accuracy of pREE when compared against mREE using indirect calorimetry was calculated using the formula: %ΔREE = ((pREE-mREE)/mREE) x 100 (37,38). Accuracy of pREE was defined as ± 10% from mREE. As indirect calorimetry measurement error is accepted at 5% (46), an error limit of ± 10% is accepted as twice the measurement error to indicate a ‘true difference’ (47,48). Underprediction of REE by the predictive equation produces a negative %ΔREE, whilst overprediction results in a positive %ΔREE.

### Metabolic Index

The metabolic index (MI) percentage was calculated using the following formula: MI = (mREE/pREE) x 100 (34,35). A metabolic index threshold of ≥ 110% was used to classify hypermetabolism (47).

## Results

### Study population

Two participants withdrew consent before indirect calorimetry was conducted. Indirect calorimetry measurements were conducted on 22 people living with MND between October 2021 and August 2022. Weight measurements were neither collected nor reported from two participants. REE could therefore not be estimated for these participants, and they were excluded from analyses. Participants who did not provide complete 24-hour urinary collections for the measurement of total urinary nitrogen were also excluded from analysis (*n* = 4). The flowchart of participant inclusion is shown in Figure 1.

**Figure 1:**
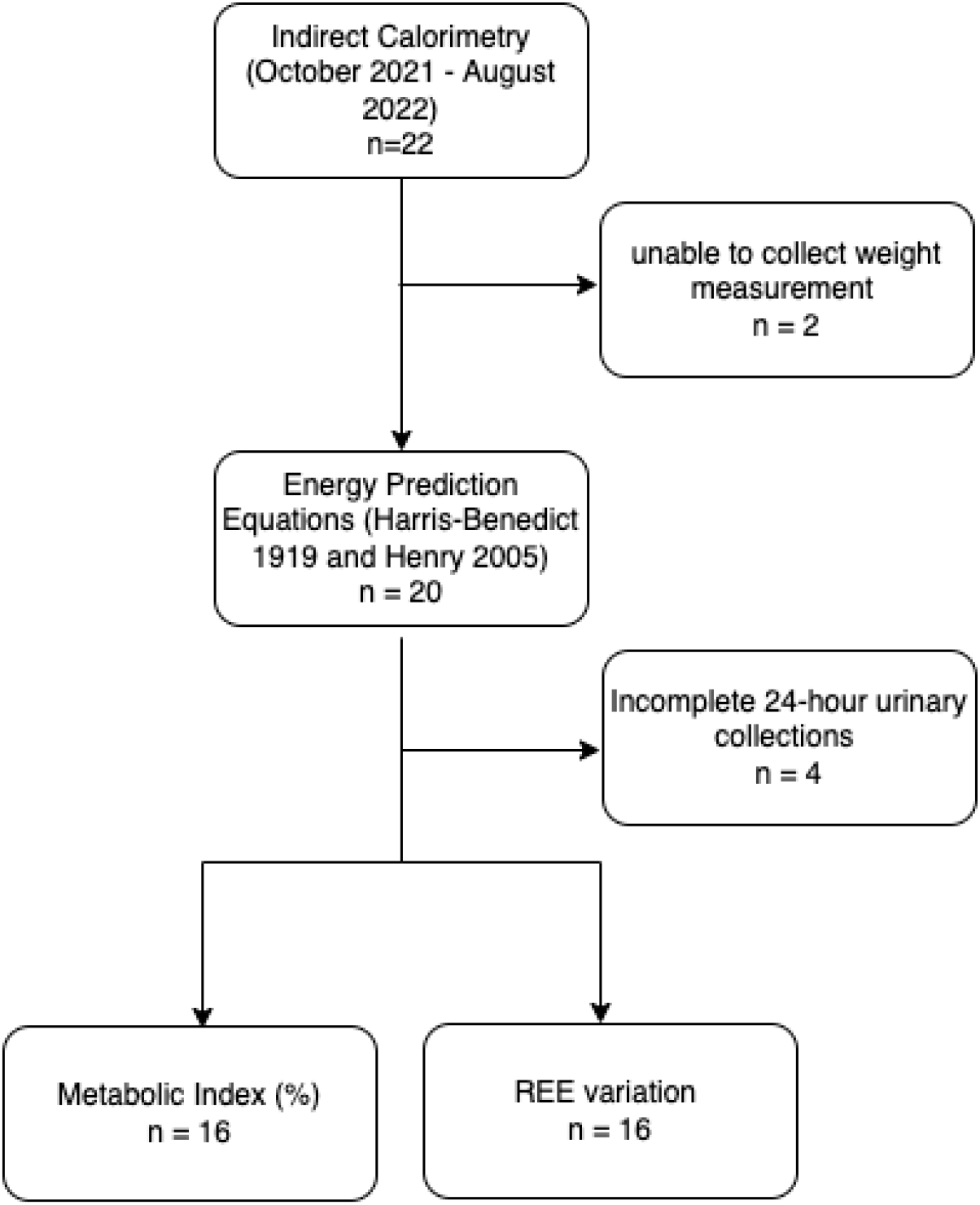
A flowchart of participants living with MND included in the study.

Of the sixteen included participants, 100% were male. Participant demographics, anthropometric measurements and disease duration from symptom onset (in months) are shown in **Table 2**.

**Table 2:** Descriptive statistics of demographic, anthropometric and clinical characteristics of participants included in the analysis (n = 16). ^a^p value of comparison between mREE and pREE. ALSFRS-R: Amyotrophic Lateral Sclerosis Functional Rating Scale (Revised); AMA: arm muscle area; BMI: body mass index; MUAC: mid upper arm circumference; mREE: measured resting energy expenditure; pREE: predicted resting energy expenditure; TSF: triceps skin fold; TUN: total urinary nitrogen; VCO_2_: volume of carbon dioxide expired; VO_2_: volume of oxygen inspired.

|  | Mean (SD) | Median (IQR) | Minimum | Maximum | *p* |
| --- | --- | --- | --- | --- | --- |
| **Age** | 62 (12.1) | 60 (55-71) | 37 | 83 |  |
| **Weight (kg)** | 81.89 (16.98) | 83.50 (70.00-91.28) | 51.40 | 117.48 |  |
| **Height (cm)** | 176.33 (6.58) | 176.15 (171.68-178.60) | 165.10 | 190.50 |  |
| **BMI (kg/m<sup>2</sup>)** | 26.14 (4.24) | 26.20 (23.48-28.26) | 17.70 | 33.00 |  |
| **Left arm AMA (cm<sup>2</sup>)** | 52.62 (14.62) | 48.66 (39.45-62.30) | 30.54 | 81.45 |  |
| **Right arm AMA (cm<sup>2</sup>)** | 52.36 (14.60) | 48.70 (42.60-62.33) | 31.84 | 81.98 |  |
| **VO<sub>2</sub> (ml/min)** | 234.05 (37.56) | 248.16 (203.30-262.42) | 163.07 | 290.73 |  |
| **VCO<sub>2</sub> (ml/min)** | 211.87 (31.36) | 219.64 (186.97-239.70) | 131.47 | 244.80 |  |
| **TUN (g/24hr)** | 11.08 (3.05) | 11.24 (8.40-12.72) | 6.69 | 17.39 |  |
| **Symptom duration (months)** | 44 (45) | 27 (18-59) | 12 | 188 |  |
| **ALSFRS-R** | 34.25 (7.04) | 31.50 (29.00-42.50) | 24.00 | 45.00 |  |
| **mREE (kcal/24hr)** | 1642 (258) | 1740 (1435-1848) | 1110 | 2015 |  |
| **Harris-Benedict (kcal/24hr)** | 1655 (265) | 1644 (1398-1819) | 1294 | 2176 | 0.87<sup>a</sup> |
| **Henry (kcal/24hr)** | 1683 (231) | 1671 (1485-1831) | 1363 | 2114 | 0.58<sup>a</sup> |

### Measured Resting Energy Expenditure

mREE was derived for each participant from the Weir equation using the volume of oxygen inspired (VO_2_), volume of carbon dioxide expired (VCO_2_) and total urinary nitrogen values (Table 2). The mean mREE for the cohort was 1642 kcal/24hr (± 258), with individual data ranging from 1110 to 2015 kcal/24hr (Figure **2**A/Table 2). This was not significantly different to the mean pREE using either the HB (1655 kcal/24hr ± 265, *p* = 0.87) or Henry (1683 kcal/24hr ± 231, *p* = 0.58) equations (Figure ***2***A/Table 2). Bivariate correlation analysis demonstrated a weak, positive relationship between mREE and both the HB (Pearson’s *r* = 0.184, *p* = 0.495) and Henry (Pearson’s *r* = 0.261, *p* = 0.329) predictive energy equations (Figure **2**B/C).

**Figure 2:**
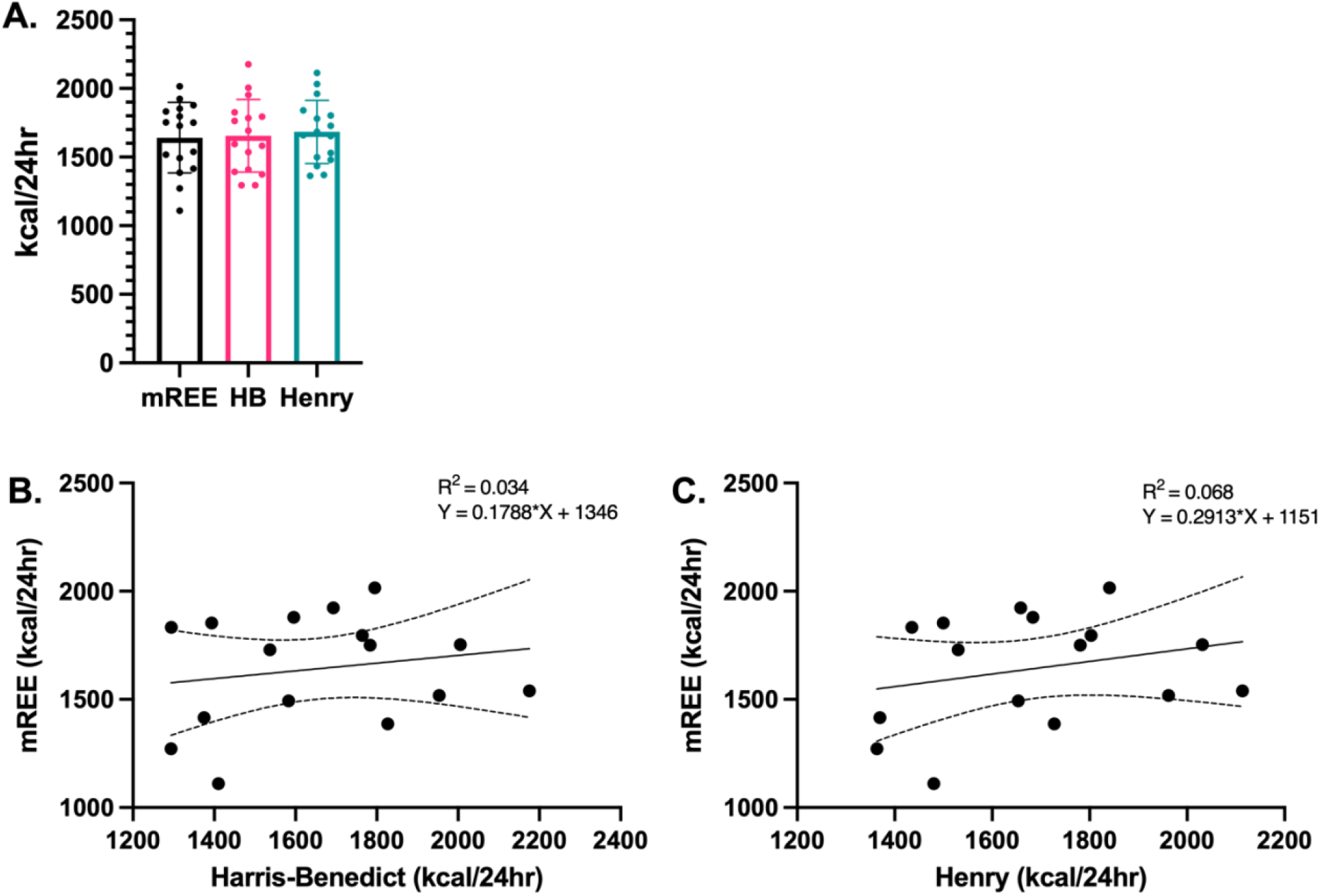
Measured and predicted resting energy expenditure (n = 16). (A) comparison of the mean ±1 SD of mREE and pREE using the HB and Henry equations. (B and C) comparison of mREE against pREE using HB and Henry equations. B and C show regression line with 95% confidence intervals. HB: Harris-Benedict; mREE: measured resting energy expenditure; pREE: predicted resting energy expenditure; SD: standard deviation.

### Metabolic Index

mREE was compared against the HB and Henry predictive equations to calculate the metabolic index for each participant. Whilst the average value for the entire cohort did not surpass the 110% threshold (mean MI = HB: 101.04% ±20.33; Henry: 98.62% ±17.40) (**Table 3**) intra-cohort analysis revealed 6/16 (37.5%) (HB) and 5/16 (31.25%) (Henry) of participants would be categorised as hypermetabolic using the 110% threshold.

**Table 3:** Metabolic index (%) = measured resting energy expenditure compared with predicted resting energy expenditure (n = 16). Derived mREE was compared against pREE using either the Harris-Benedict or Henry equation to calculate the metabolic index (%). Hypermetabolism is indicated using a metabolic index threshold of 110%. MI: metabolic index.

|  | Mean (SD) | Median (IQR) | Minimum | Maximum | Hypermetabolic participants n/N (%) |
| --- | --- | --- | --- | --- | --- |
| **pREE Harris-Benedict MI (%)** | 101.04 (20.33) | 100.06 (80.90-113.32) | 70.75 | 141.58 | 6/16 (37.5) |
| **pREE Henry MI (%)** | 98.62 (17.40) | 98.93 (81.77-112.65) | 72.82 | 127.69 | 5/16 (31.3) |

### REE Variation

Whilst a weak, positive relationship between either the HB or Henry equation and mREE exists in our cohort (**Figure 2**), correlation analysis only presents the linear relationship between two variables, but not agreement (49). Bland-Altman limits of agreement analysis helped visualise bias and precision between the measured and predicted REE (**Figure 3**).

**Figure 3:**
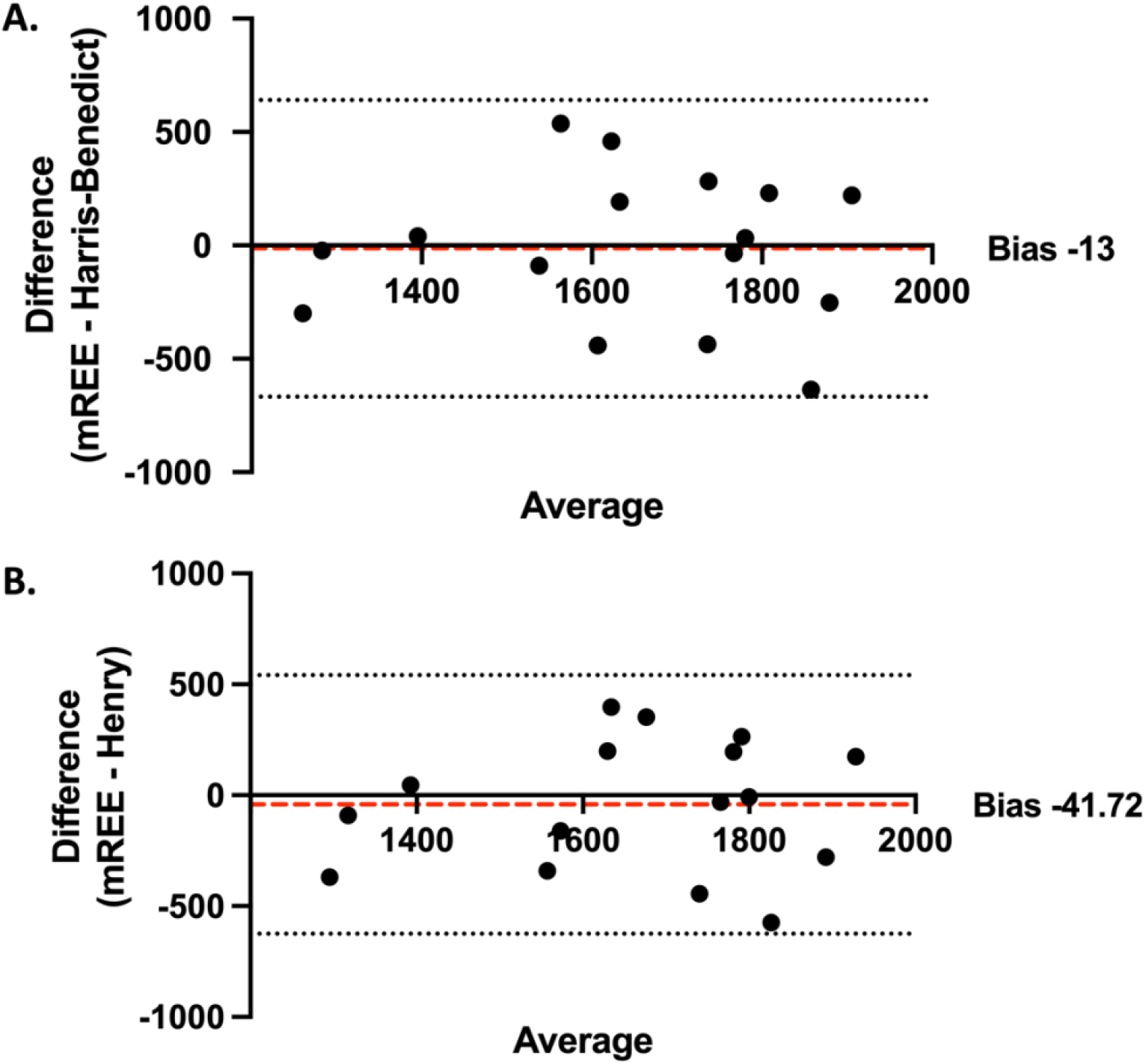
Agreement between measured and predicted resting energy expenditure (kcal/day) (n = 16). (A) HB 1919. (B): Henry 2005. The mean bias between mREE and pREE is indicated by a red dotted line, with 95% CIs indicated by black dotted lines. CI: confidence intervals; HB: Harris-Benedict; mREE: measured resting energy expenditure, pREE: predicted resting energy expenditure.

On average, both the HB and Henry equations overestimated REE when compared to mREE (mean bias: HB = -13 kcal/day; Henry = -41.72 kcal/day) (**Figure *3***A/B). However, a high degree of variability was observed within the cohort, with both predictive equations not only overestimating, but also underestimating, REE (HB: range = -538 -636 kcal/day, 95% CI = - 667–641 kcal/day; Henry: range = -397 - 575 kcal/day, 95% CI = -624.8–541.3 kcal/day) (**Figure 3**).

When assessed for REE variation (%ΔREE), pREE by both the HB and Henry equations accurately reflected mREE at group level (± 10%) (mean %ΔREE = HB: 2.81 ± 20.81; Henry: 4.51% ± 18.98) (**Table 4**). However, inter-individual %ΔREE analysis within this cohort revealed both the HB and Henry equations inaccurately reflected mREE for 68.7% of participants (**Table 4**).

**Table 4:** Resting energy expenditure variation: percentage difference between measured and predicted resting energy expenditure (n = 16). %ΔREE: percentage difference between measured and predicted resting energy expenditure.

|  | Mean (SD) | Median (IQR) | Minimum | Maximum | Accurate<br>n/N (%) |
| --- | --- | --- | --- | --- | --- |
| **pREE Harris-Benedict (%ΔREE)** | 2.81 (20.81) | -0.03 (-11.75-23.86) | -29.37 | 41.34 | 5/16 (31.3) |
| **pREE Henry (%ΔREE)** | 4.51 (18.98) | 1.08 (-11.23-22.41) | -21.68 | 37.33 | 5/16 (31.3) |

To determine factors influencing over- or under-estimation of REE, independent variables forming both predictive equations, such as age, sex, weight and height were compared against the %ΔREE for participants in this cohort (**Figure 4**).

**Figure 4:**
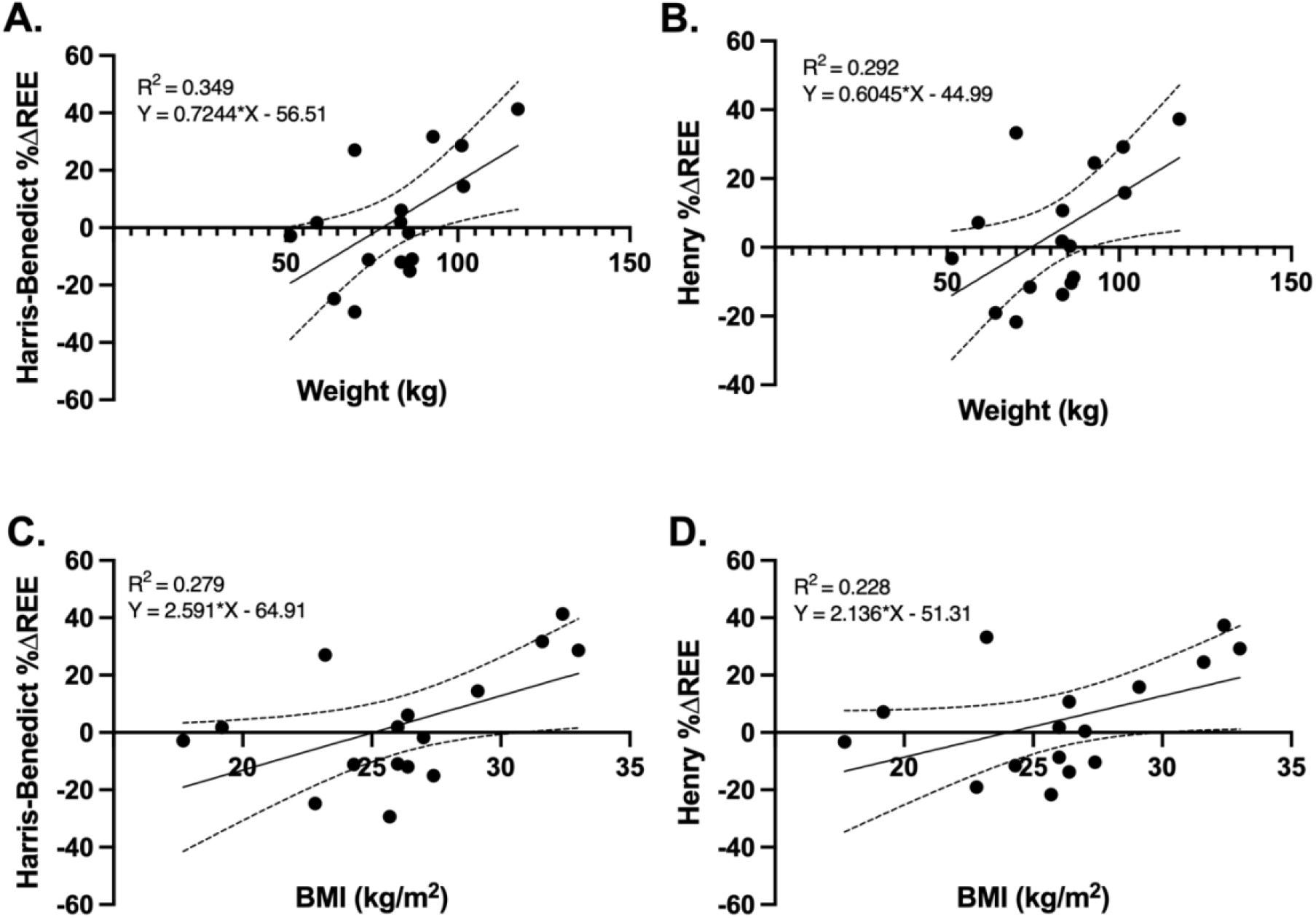
Comparing weight and BMI to percentage difference between measured and predicted resting energy expenditure (n = 16). (A/B) Weight (kg) against the %ΔREE using pREE by HB and Henry, respectively. (C and D) BMI (kg/m^2^) against the %ΔREE using pREE by HB and Henry equations. HB: Harris-Benedict; pREE: predicted resting energy expenditure; %ΔREE: percentage difference between measured and predicted resting energy expenditure.

%ΔREE was significantly strongly, positively correlated with weight for both the HB and Henry equation (HB: *r* = 0.591, *p* = 0.016; Henry: *r* = 0.541, *p* = 0.031) (**Figure 4**A/B). As weight is a constituent element of BMI, a similar relationship was expected between BMI and %ΔREE for both equations; but this was significant only for the HB equation (HB: *r* = 0.528, *p* = 0.035; Henry: *r* = 0.477, *p* = 0.061) (**Figure 4**C/D). These correlations demonstrated that both the HB and Henry equations overpredicted REE (a positive %ΔREE) in heavier individuals, but underpredicted REE (a negative %ΔREE) in lighter individuals, when compared with mREE.

There was no correlation between arm muscle area, a proxy for lean body mass, and %ΔREE (HB: *r* = 0.111, *p* = 0.682; Henry: *r* = 0.049, *p* = 0.857).

### Does REE variation influence the metabolic index?

Hence, we raised the question as to whether an underprediction of REE using predictive energy equations, when compared against mREE, also biases the identification of hypermetabolism, using the 110% metabolic index threshold.

The %ΔREE was significantly negatively correlated with the metabolic index for both the HB and Henry equations (HB: *p* = -0.979, *r* = <0.001; Henry *p* = 0.988, *r* = <0.001) **(Figure 5)**. Hence, the lower the %ΔREE (i.e., the greater the underprediction by pREE) the greater the metabolic index.

**Figure 5:**
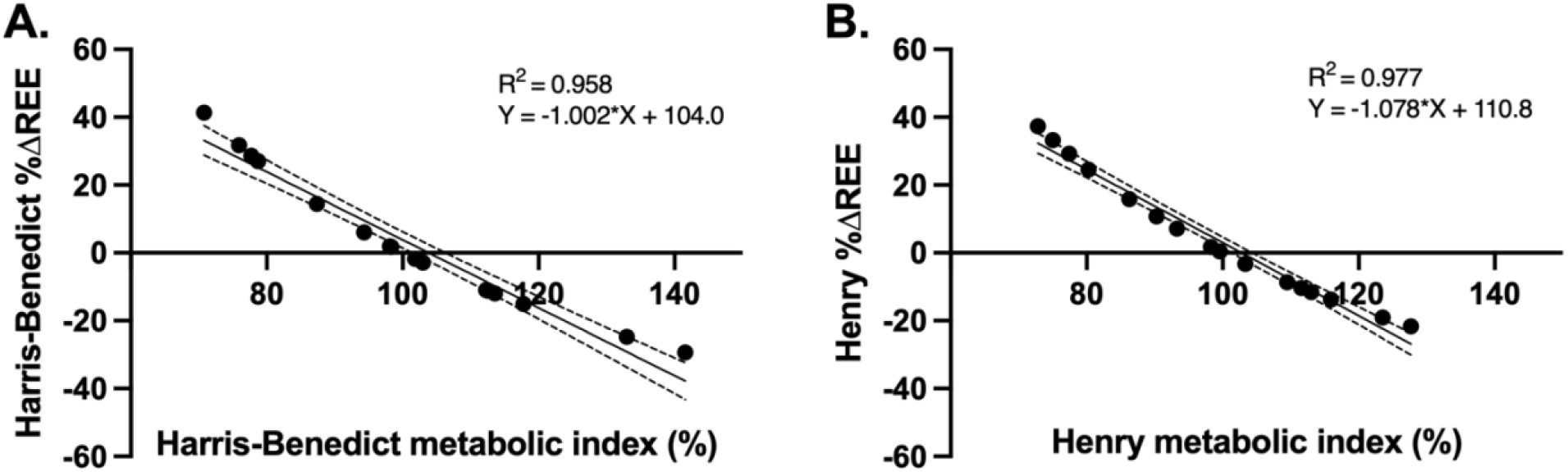
Comparing resting energy expenditure variation against the metabolic index. %ΔREE against the metabolic index using HB (panel A) and Henry (panel B) equations. HB: Harris-Benedict; MI: metabolic index; %ΔREE: percentage difference between measured and predicted resting energy expenditure.

### kcal/kg/day

Four participants (25% of the study population) were excluded from analysis. pREE was found to be 1798 kcal/24hrs (± 249). This was not significant when compared against mREE in the same individuals (mean mREE: 1701 ± 272 kcal/24hr; *p* = 0.290). When assessed for accuracy, the mean REE variation using kcal/kg/day was 8.00%, and was found to be accurate in 7/12 (58.3%) of participants. The average metabolic index was 95.64%, with 1/12 (8.33%) participants surpassing the metabolic threshold of 110%.

## Discussion

In our study, mREE derived by means of indirect calorimetry and total urinary nitrogen analysis was similar to previous research findings in plwMND (32,33,36,37,50). mREE was compared to pREE using HB, Henry and kcal/kg/day equations to critically evaluate the suitability of using the metabolic index to indicate hypermetabolism in plwMND.

More than 100 predictive energy equations exist, which presume a linear relationship between REE and independent variables such as age, weight, height, and other body composition indices (51). A potential disadvantage of predictive energy equations is that they are predominantly derived from young, healthy, Caucasian individuals; hence, may not accurately reflect mREE in critical, chronic illness (52) or MND patients (36–38,53).

### What is the importance of identifying hypermetabolism?

Hypermetabolism has been shown to be a prognostic indicator of survival, functional change and weight loss (8,17,18,53–56). Therefore, it is important to identify hypermetabolism in individuals to optimise nutritional management. The metabolic index is not a readily-accessible tool that can be calculated by dietitians, and there is a need for MND-specific predictive equations that could be applied to inform appropriate dietetic nutritional management and incorporate a metabolic component.

### Predictive energy equations overestimate resting energy expenditure at group level

The inclusion of the Henry equation in this study was informed by the results of a recent large UK-wide survey of dietetic practice in MND (22). The accuracy of the Henry equation when compared to mREE has not been previously assessed in MND. We found the Henry equation to accurately reflect mREE in 5/16 (31.3%) of our participants. This is the same as for the HB equation (**Table 4**). In line with previous MND literature utilising the HB equation to calculate pREE (37,38,57), our results show an overall overestimation of pREE with a lack of precision, as demonstrated by the high confidence intervals (**Figure 3**). This intra-cohort variability resulted in an underestimation of energy requirements by up to 636 kcal/24hr, as well as overestimation by 538 kcal/24hr (**Figure 3**). To explain this variability, we conducted bivariate correlations of continuous variables incorporated within both the HB and Henry predictive equations (i.e., weight, height and age) against mREE.

### Weight informs predictions of resting energy expenditure

We have shown that weight correlates in a linear relationship with REE variation (51) (**Figure 4**A/B). This suggests that the lighter the individual, the greater the underestimation of pREE, producing a lower, negative %ΔREE, and vice versa (**Figure 4**). The clinical implications of this inaccuracy could result in under-or over-feeding patients with potentially detrimental clinical outcomes. Underfeeding would be more likely to occur in lighter individuals, contributing towards accelerated muscle wastage, malnutrition and irreversible weight loss. Conversely, a potential consequence of overfeeding (caused by an overestimation of pREE in heavier individuals) is hypercapnia (58), which can cause respiratory acidosis, inducing further respiratory implications (59).

Weight measurements represent lean and fat mass, both of which have different contributions to REE. Whilst lean body mass (including visceral organs and skeletal muscle) is highly metabolically active, fat mass (such as adipose tissue) is largely metabolically inactive (19,34,60). The reduction of skeletal muscle in plwMND deviates from the underlying assumed metabolic contributions that are observed in healthy individuals, altering REE (29,61–64). This may explain the overestimation of pREE at group level. It has been suggested that predictive equations may have increased accuracy if pre-morbid body weight was used, in place of current body weight (51).

### The metabolic index does not appropriately identify hypermetabolism

The inaccuracy of predictive energy equations led us to question the suitability of applying these equations to calculate the metabolic index as a method to identify hypermetabolism in MND. We have shown that the metabolic index calculated using the HB, Henry and kcal/kg/day equations in this cohort does not indicate hypermetabolism at group level (mean metabolic index range: 95.64-101.04%) (**Table 3**). However, when analysed individually, up to one third of our study population were found to be hypermetabolic (**Table 3**). More pertinently, the number of individuals identified as hypermetabolic was dependent on the predictive equation used, i.e., 37.5 % when calculated using the HB equation, 31.3 % when using the Henry and 8.33% when using kcal/kg/day.

### Current recommendations in MND dietetic practice

The conceptualisation of predictive energy equation inaccuracy in MND is not a novel one (37,38). Whilst additional MND-specific predictive equations have been developed in this knowledge (37,40,65), these require body composition measurements such as bioelectric impedance analysis (BIA). A potential limitation of this is the inclusion of additional predictive equations within BIA analysis, which may further compound measurement error (66,67).

UK MND dietetic practice is informed by guidelines released by the British Dietetic Association (BDA) Parental and Enteral Nutrition Group (PENG). These guidelines currently recommend estimation of REE using 22-24 kcal/kg/day for plwMND (40). This calculation was devised from mREE using indirect calorimetry conducted in two cohorts of plwMND (34,65). One weakness of this approach however, is that this equation has only been validated in MND for individuals with a BMI indicating healthy or normal weight and overweight (18.5-30.0 kg/m^2^), and it may not be appropriate for individuals with BMI extremes (68). When analysed in an appropriate sub-group from our cohort (12/16 (75%) of the original study population), one individual surpassed the metabolic index threshold of 110%. pREE using kcal/kg/day was also underestimated in the same individual, and most notably, this individual fell below the 25^th^ percentile for weight (kg) in this cohort. This reinforces the data presented in this article, suggesting the lighter the individual, the greater the underprediction of REE, which biases towards a metabolic index ≥ 110%.

### What does this mean for future research?

It would be easy to conclude that the HB, Henry and kcal/kg/day equations are unsuitable for estimating energy requirements in all individuals living with MND. However, these equations were accurate in 31.3-58.3% of participants within this cohort. It might therefore be more appropriate to develop weight or BMI guidance ranges for when these equations may appropriately reflect mREE. Application of predictive equations to individuals outwith these weight or BMI ranges would need to be utilised with caution.

### Considerations

Undertaking research with this frail and often mobility restricted cohort of patients does not come without practical challenges, and researchers often have to take a pragmatic approach. Although we were able to detect statistically significant relationships, the sample size and lack of gender diversity limits our ability to draw firm conclusions for the wider MND population. Out of the 22 initially recruited participants, 16 were included in the analysis because of challenges around obtaining valid weight measurements and complete 24hr urinary collections (**Figure 1**). The all-male sample may be a result of the requirement of a 24hr urinary sample collection, which may have deterred the participation of female patients. To reduce participation burden, participants were not required to fast before indirect calorimetry measurement, and we acknowledge the likelihood of a component of dietary-induced thermogenesis within the obtained indirect calorimetry measurements. Intra-cohort variation in our sample was such that it was not meaningful to stratify individuals according to clinical characteristics, e.g., functional status, duration of disease from onset of symptoms, or disease severity (**Table 2**) in order to examine relationships between clinical characteristics and mREE. This research was designed as an exploratory pilot study, and as such we did not measure all possible confounders that may contribute towards REE (e.g., body temperature (69)).

## Conclusion

Although our cohort was not hypermetabolic as a group, intra-cohort analysis revealed high variations and inaccuracies when using either the HB, Henry or kcal/kg/day predictive energy equations to estimate REE. Weight and BMI appear to be an important contributing factor to the under-or over-prediction of REE, e.g., the lighter the individual, the greater the underprediction of REE using either the HB or Henry equation. The %ΔREE appears to negatively correlate with the metabolic index, whereby the greater the underprediction of REE, the greater the metabolic index. This subsequently biases the classification of hypermetabolism towards individuals who are lighter. We suggest this %ΔREE is more likely to be attributed to the assumed metabolic contributions from a given weight included in the predictive energy equations, rather than resembling a true clinically significant raised REE.

## Data Availability

All data produced in the present work are contained in the manuscript

## Acknowledgements

With thanks to the NIHR Short Placement Award for Research Collaboration (SPARC) for support in this research, as well as the Advanced Wellbeing Research Centre, Sheffield Hallam University for technical support, indirect calorimetry equipment and supporting infrastructure.

## Abbreviations

**Full wording**

ALS: Amyotrophic Lateral Sclerosis
AMA: Arm Muscle Area
BIA: Bioelectrical Impedance Analysis
BMI: Body Mass Index
CI: Confidence Interval
DEXA: Dual Energy X-Ray Absorptiometry
Fe: Fraction of expired
Fi: Fraction of inspired
HB: Harris-Benedict
IC: Indirect Calorimetry
mREE: Measured Resting Energy Expenditure
MI: Metabolic Index
MND: Motor Neuron Disease
MUAC: Mid Upper Arm Circumference
pREE: Predicted Resting Energy Expenditure
REE: Resting Energy Expenditure
SD: Standard Deviation
TSF: Triceps Skin Fold
VO_2_: Volume of Oxygen inspired
VCO_2_: Volume of Carbon Dioxide expired
%ΔREE: Percentage Difference in REE
24hr: Twenty-four hour

